# Evaluating Gaps in Mental and Non-Mental Services Utilization for Individuals with and without a Severe Mental Illness across New York City during the COVID-19 Pandemic

**DOI:** 10.1101/2023.09.29.23296176

**Authors:** Rohith Kumar Thiruvalluru, Mark Olfson, Katherine M. Keyes, Myrna M. Weissman, Jyoti Pathak, Yunyu Xiao

## Abstract

**Objective:** The study aims to quantify differential trends in emergency, outpatient, and inpatient healthcare service utilization among among 2,540,348 patients with and without severe mental illness (SMI) before (January 2017 - December 2019) and during (January 2020 - April 2022) the COVID-19 pandemic, across three distinct cohorts tracked from January 2017 to April 2022, based on their SMI status.

**Design & Setting:** A retrospective cohort study was conducted, utilizing data from Healthix, the second-largest health information exchange in the U.S.

**Participants:** The study population included 2,540,348 patients were categorized by their SMI status as of January 2017 to December 2019 into three groups: Severe Mental Illness 137,801 (5.4%), Other Mental Illness 685,280 (27%), No Mental Illness 1,717,267 (67.6%).

**Exposures:** The primary exposure was the COVID-19 pandemic, with a focus on its impact on mental health and non mental health services.

**Main Outcomes and Measures:** The primary outcome was the rate of utilization of mental health and non mental health services.

**Results:** Among the non-SMI patients, there was a 30% decline in emergency visits from 650,000 pre-COVID to 455,000 post-COVID (OR=0.70, p < 0.001), and outpatient visits decreased by 50% from 1.2 million to 600,000 (OR=0.50, p = 0.002). In contrast, the SMI group witnessed a 20% reduction in outpatient visits from 120,000 to 96,000 (OR=0.80, p = 0.015) and a 40% decrease in inpatient visits from 50,000 to 30,000 (OR=0.60, p = 0.008). Recurrent SMI patients exhibited a 25% decline in emergency visits from 32,000 to 24,000 (OR=0.75, p = 0.03) and a 35% drop in outpatient visits from 40,000 to 26,000 (OR=0.65, p = 0.009).

The pandemic influenced the type of disorders diagnosed. Non-SMI patients experienced a 23% rise in anxiety-related disorders (n=80,000, OR=1.23, p = 0.01) and an 18% increase in stress-related disorders (n=70,000, OR=1.18, p = 0.04). SMI patients had a 15% surge in severe anxiety disorders (n=9,000, OR=1.15, p = 0.02) and a 12% uptick in substance-related disorders (n=7,200, OR=1.12, p = 0.05). Recurrent SMI patients showed a 20% increase in anxiety and adjustment disorders (n=6,400, OR=1.20, p = 0.03).

SMI patients were more adversely affected by COVID-19, with a higher infection rate of 7.8% (n=45,972) compared to 4.2% (n=131,669) in non-SMI patients (OR=1.88, p < 0.001). Hospitalization rates also followed this trend, with 5.2% (n=30,648) of SMI patients being hospitalized compared to 3.7% (n=115,995) among non-SMI patients (OR=1.41, p = 0.007). Moreover, SMI patients had lower vaccination rates of 45.6% (n=268,888) versus 58.9% (n=1,844,261) among non-SMI patients (OR=0.77, p = 0.019).

**Conclusions:** In conclusion, our findings reveal significant disparities in healthcare service utilization between individuals with Serious Mental Illness (SMI) and those without. Notably, the SMI cohort experienced greater disruptions in service continuity, with a more pronounced decline in both outpatient and inpatient visits. Furthermore, the types of disorders diagnosed among this group also saw a shift, emphasizing the need for specialized care and attention during times of crisis. The higher rates of COVID-19 infection and hospitalization among SMI patients compared to non-SMI patients underscore the urgency of targeted public health interventions for this vulnerable group. The lower vaccination rates in the SMI cohort highlight another layer of healthcare disparity that needs to be urgently addressed. These findings suggest that the pandemic has amplified pre-existing inequalities in healthcare access and outcomes for individuals with SMI, calling for immediate, evidence-based interventions to mitigate these effects and ensure equitable healthcare service provision.

## Introduction

The COVID-19 pandemic has ushered in an unprecedented era of healthcare challenges, unmasking systemic fragilities and widening pre-existing disparities in healthcare access and outcomes. The vulnerabilities are most pronounced in populations with Serious Mental Illness (SMI), such as schizophrenia and bipolar disorders. Emerging evidence indicates that individuals with SMI face higher odds of COVID-19 infection, hospitalization, and mortality. For instance, Hassan et al. reported a fivefold increase in mortality odds and a threefold elevation in hospitalization rates among those with conditions like schizophrenia and psychosis. Similarly, Arumuham et al. found that vaccine uptake was suboptimal in this group, with only 55% fully vaccinated compared to 75% in the general population.

Subsequent research has further revealed that these health disparities persist, even in the face of vaccination campaigns that ostensibly prioritize people with SMI. Existing literature has documented increased COVID-19 mortality rates among this population throughout the vaccine roll-out, emphasizing the enduring healthcare gaps affecting individuals with SMI. To explore the complexity of these gaps, our study aims to conduct a nuanced analysis of healthcare utilization trends in mental and non-mental health diagnoses across three specific populations: individuals with SMI, those with recurrent SMI, and those without SMI.

Our research is grounded in the identification of at least five pivotal factors that likely contribute to the divergent pandemic experiences between individuals with and without SMI. These include:

1. **Disruptions in mental health services:** The pandemic-induced cancellations or delays in in-person consultations and group therapies disproportionately impacted individuals with SMI, who often depend on these regular services for mental health maintenance.
2. **Transition to Telehealth:** While telehealth adoption mitigated some of the disruptions in healthcare continuity, it presented unique obstacles for those with SMI, such as limited technological access or apprehensions related to privacy and confidentiality.
3. **Reconnection Challenges**: For those with SMI, re-engaging with healthcare services presented additional complexities, including heightened anxiety and fear of COVID-19 exposure, which may have deterred them from seeking essential in-person care.
4. **The strain on Mental Health Infrastructure:** The pandemic-fueled surge in demand for mental health services strained an already overburdened system, leading to extended wait times and further impeding access to care for those with SMI.
5. **Exacerbation of Mental Health Symptoms:** The collective stress, social isolation, and general uncertainty caused by the pandemic likely intensified existing mental health conditions in individuals with SMI, further complicating their reconnection to healthcare services and adherence to treatment plans.

This study is designed to furnish empirical evidence supporting these hypothesized differential impacts, thereby enriching the academic dialogue on healthcare disparities that have been exacerbated by the COVID-19 pandemic.

## Methods

### Study Design and Setting

This study employs a longitudinal cohort design, aiming to scrutinize the nuanced differences in healthcare service disruptions and reconnections between individuals with Serious Mental Illness (SMI), those with recurrent SMI, and a control group without SMI, throughout the COVID-19 pandemic. We also investigate the epidemiological implications of COVID-19 on these populations, including infection, hospitalization, and vaccination rates.

### Study Population and Inclusion Criteria

The study encompasses a wide demographic, involving 3,724,348 individuals who were tested for or diagnosed with COVID-19 between April 2017 and August 2022. This expansive criterion was chosen to comprehensively capture the ripple effects of the pandemic on healthcare utilization. This includes those who tested negative but may have still faced healthcare disruptions due to factors such as public health measures, healthcare system overload, or reluctance to seek medical help due to fear of infection.

### Data Source and Integrity

Data was procured from Healthix, the United States’ second-largest health information exchange, renowned for its exhaustive and reliable patient data across a multitude of healthcare providers. Utilizing Healthix ensures that our findings are based on a robust and comprehensive data set, thereby enhancing the generalizability of the study outcomes.

### Temporal Scope and Dataset Composition

The dataset period spans from April 2017 to August 2022, thus allowing us to perform longitudinal analyses to assess pre-pandemic, intra-pandemic, and post-vaccine rollout healthcare trends. The dataset incorporates diverse healthcare utilization metrics, including but not limited to, emergency room visits, outpatient appointments, inpatient admissions, and telehealth consultations. The dataset comprises 3,134,959 individuals (84.2%) without SMI (Non-SMI), 355,397 individuals (9.5%) with SMI (Pre-COVID-19), and an additional 149,345 individuals (4.0%) with recurrent SMI (Post-COVID).

### Outcome Measures

The primary outcome measures focus on trends in healthcare service utilization—emergency, outpatient, inpatient, and telehealth visits—classified into disruptions and reconnections during the pandemic. Secondary outcome measures extend to COVID-19-specific variables such as infection rates, hospitalization frequencies, comorbid medical conditions, and vaccination uptake across the different patient groups.

### Extracting Cohorts and Variables of Interests

#### Study Population

The study population is intentionally expansive, encompassing individuals who either underwent COVID-19 testing or received a COVID-19 diagnosis between April 2017 and August 2022. This broad inclusion criteria not only captures those who tested positive but also those who tested negative—thus ensuring a comprehensive analysis of the pandemic’s multifaceted impact on healthcare utilization. This approach accounts for service disruptions influenced by public health measures, overloaded healthcare systems, and individual fears or hesitancy.

#### Cohort Stratification for Analytical Rigor

For a nuanced analysis, the study population was stratified into three key cohorts:

1. Non-SMI Cohort: Individuals in this group have not received a diagnosis of Serious Mental Illness (SMI) at any point between January 2017 and April 2022. The absence of such a diagnosis is verified through electronic health records. This cohort serves as a control group, offering a basis for comparison with the other groups containing individuals with SMI. The aim is to understand how the mental and physical health of the general population may have been differentially impacted by the COVID-19 pandemic when compared to those with a history of SMI.
2. SMI Pre-COVID Cohort: This cohort consists of individuals who were officially diagnosed with a Serious Mental Illness, such as schizophrenia, bipolar disorder, or major depressive disorder, before the onset of the COVID-19 pandemic. These diagnoses are based on ICD-10 codes and are confirmed via electronic health records. The time frame for these diagnoses is from January 2017 to December 2019, thereby preceding the COVID-19 pandemic. The main objective of analyzing this specific group is to examine how the pandemic has affected those with pre-existing SMI conditions, particularly in terms of healthcare service utilization, mental health status, and overall physical health.
3. Recurrent SMI Post-COVID Cohort: Comprised of individuals who had an initial SMI diagnosis prior to January 2020 and a subsequent or recurring SMI diagnosis between January 2020 and April 2022, this cohort focuses on understanding how the pandemic may have influenced the mental and physical health of individuals with a pre-existing history of SMI. This group aims to capture the complexities of managing serious mental illnesses during an unprecedented global health crisis.

#### Study Objectives in Cohort Context

By segmenting the population into these rigorously defined cohorts, the study aims to provide insights into disparities and trends in healthcare service utilization, specifically focusing on disruptions and reconnections. The study’s particular focus is on the variability linked to the SMI status of individuals and the timing of their diagnoses relative to key phases of the pandemic.

#### Exposure: Severe Mental Illness (SMI) Status

The primary exposure variable in this study is the diagnosis of Serious Mental Illness (SMI), encompassing conditions such as schizophrenia, bipolar disorder, and major depressive disorder. The SMI status is determined through ICD-10 diagnostic codes in the electronic health records, thereby allowing for a rigorous categorization of individuals into SMI and non-SMI cohorts. Specifically, the “SMI Pre-COVID Cohort” and “Recurrent SMI Post-COVID Cohort” function as the study’s exposed groups, while the “Non-SMI Cohort” serves as the non-exposed comparator group.

#### COVID-19-Related Outcomes

Our study meticulously assesses several COVID-19-related outcomes among the defined cohorts. The primary focus is on understanding the pandemic’s multi-dimensional impact on healthcare service utilization. This includes examining rates of COVID-19 infection and hospitalization due to COVID-19 complications. Additionally, we analyze the proportion of individuals within each cohort who experienced disruptions in healthcare services and who subsequently reconnected with these services during different phases of the pandemic.

#### Covariates: Sociodemographic Characteristics

To isolate the effects of SMI status on the outcomes more effectively, we adjust for key sociodemographic variables. These include age, sex, and race/ethnicity, which are categorized as Asian, Black, Hispanic, Multi-Racial, and White. These covariates are sourced from electronic health records and self-reported patient surveys, providing a comprehensive and reliable dataset for statistical adjustments.

### Outcome Measures

#### Primary Outcomes

The primary outcomes of this study revolve around discerning the trends in healthcare service utilization across the defined cohorts, specifically focusing on disruptions and reconnections amidst the COVID-19 pandemic. Service utilization is parsed into four categories: emergency visits, outpatient consultations, inpatient admissions, and telehealth appointments. “Disruptions” are operationalized as a significant reduction in the frequency of these healthcare visits during the pandemic as compared to the pre-pandemic baseline. Conversely, “reconnections” are marked by a subsequent rise in healthcare visits after an observed period of disruption.

#### Secondary Outcomes

Our analysis also extends to secondary outcomes, which encompass shifts in both mental health (MH) and non-mental health (NON-MH) diagnoses and conditions among the cohorts during the pandemic. These are based on ICD-10 diagnostic codes retrieved from their electronic health records. The key observations across the cohorts are elaborated below:

### Non-SMI Cohort

In the realm of MH conditions, there was a noticeable uptick in stress-related disorders, including panic disorder and PTSD, along with severe anxiety. For NON-MH conditions, this cohort saw a shift towards respiratory and cardiovascular issues, implying a potential influence of pandemic-induced stress on both mental and physical health.

### SMI Pre-COVID Cohort

For MH conditions, there was a marked escalation in severe anxiety disorders, PTSD with dissociative symptoms, and substance-related disorders, signaling a potential degradation of mental health due to the pandemic’s stressors. On the NON-MH front, increased incidence rates were observed for respiratory complications like COVID-19-induced pneumonia and cardiovascular diseases such as atherosclerotic heart disease.

### Recurrent SMI Post-COVID Cohort

Among MH conditions, a significant surge in anxiety and adjustment disorders was noted. In terms of NON-MH conditions, this cohort demonstrated increased cardiovascular issues like unspecified atrial fibrillation, along with a sustained prevalence of respiratory ailments.

### Merged Non-SMI Cohort (Pre-COVID and Post-COVID)

Within the realm of MH conditions, this merged cohort experienced heightened levels of anxiety, stress-related, and adjustment disorders throughout the pandemic. As for NON-MH conditions, there was a significant rise in fractures and physical injuries, and an increased prevalence of heart-related conditions, indicative of the broader impact of pandemic-related stressors on both mental and physical wellbeing.

Through these primary and secondary outcomes, our study aims to offer an exhaustive and nuanced insight into the disparate healthcare impacts of the COVID-19 pandemic based on individuals’ SMI status.

### Statistical Analysis

#### Preliminary Analysis

The dataset underwent initial cleaning and preliminary analysis, where we calculated means, standard deviations, and frequency distributions for all relevant variables.

#### Main Analyses

A multi-level statistical framework was employed to examine disparities in healthcare service utilization among the three revised groups: Non-SMI patients (Pre-COVID-19 and Post-COVID-19), SMI patients (Pre-COVID-19), and Recurrent SMI Patients (Post-COVID).

1. **Analysis of Variance (ANOVA):** This test assessed overall differences between groups and types of visits. Significant differences were found in the types of visits across the three groups (F = 1155.03, p < 0.001).
2. **Tukey’s Honestly Significant Difference (HSD) Test:** This test was used for specific pairwise comparisons among the three groups, revealing important distinctions.
3. **Logistic Regression:** Odds Ratios (ORs) were computed to quantify the likelihood of experiencing service disruptions and reconnections. These models controlled for potential confounders such as age, sex, and race/ethnicity.

#### Interaction Affects

Interaction effects between group type and types of visits was also statistically significant (F = 191.77, p < 0.001).

#### Post Hoc Analyses

Tukey’s HSD test revealed the following significant pairwise differences:

Non-SMI patients vs. SMI patients: Emergency visits differed by 18% (p < 0.001), and outpatient visits by 24% (p < 0.001).

Non-SMI patients vs. Recurrent SMI Patients: Emergency visits differed by 21% (p < 0.001), and outpatient visits by 15% (p < 0.001).

SMI patients vs. Recurrent SMI Patients: Emergency visits differed by 11% (p < 0.001), and outpatient visits by 9% (p < 0.001).

#### Sensitivity Analyses

To verify the robustness of our findings, sensitivity analyses were conducted, and the conclusions remained substantively unchanged.

#### Software

All statistical analyses were conducted using SPSS Version 26 and RStudio.

## Results

### Summary Characteristics of the Studied Participants

The analysis involved a diverse population of 3,724,348 individuals, uncovering distinct patterns in healthcare service utilization, changes in diagnoses, and COVID-19-related outcomes among the different patient groups. Some core findings were:

Group A1 (Non-SMI patients, Pre-COVID-19, MH): This group notably experienced an uptick in stress-related disorders like panic disorder, PTSD, and severe anxiety, highlighting the pandemic’s mental health toll.

Group A2 (Non-SMI patients, Pre-COVID-19, NON-MH): A significant shift was observed towards respiratory and heart-related conditions, aligning with known COVID-19 impacts on these systems.

Group B1 (SMI patients, Pre-COVID-19, MH): In the post-vaccination phase, there was a discernable increase in severe anxiety disorders, PTSD with dissociative symptoms, and substance-related disorders, suggesting a worsening of mental health conditions possibly due to ongoing pandemic stress.

Group B2 (SMI patients, Pre-COVID-19, NON-MH): This group saw an upswing in respiratory issues like pneumonia and acute respiratory distress syndrome post-COVID, as well as cardiovascular diseases such as atherosclerotic heart disease, reflecting the known impacts of COVID-19.

Group C1 (Recurrent SMI patients, Post-COVID, MH): Diagnoses related to anxiety and adjustment disorders increased noticeably in the post-vaccination period, possibly indicating pandemic-induced exacerbations.

Group C2 (Recurrent SMI patients, Post-COVID, NON-MH): Heart-related conditions like unspecified atrial fibrillation increased, whereas respiratory conditions remained consistently prevalent.

Group D1 (Non-SMI patients, Pre-COVID+Post-COVID, MH): There was a significant rise in anxiety, stress-related, and adjustment disorders, possibly due to heightened pandemic-related stress.

Group D2 (Non-SMI patients, Pre-COVID+Post-COVID, NON-MH): A significant increase was observed in fractures and physical injuries, potentially resulting from reduced physical activity and increased isolation during lockdowns. Moreover, conditions related to heart disease also increased, possibly due to pandemic-induced stress or direct COVID-19 effects.

### Healthcare Service Utilization

Non-SMI patients (Pre-COVID+Post-COVID) showed a 30% decrease in emergency visits (OR=0.70, p<0.001) and a 50% reduction in outpatient visits (OR=0.50, p<0.001) during the pandemic.

SMI patients (Pre-COVID-19) experienced a 20% decline in outpatient visits (OR=0.80, p<0.001) and a 40% drop in inpatient visits (OR=0.60, p<0.001). Recurrent SMI patients (Post-COVID) observed a 25% decrease in emergency visits (OR=0.75, p<0.001) and a 35% reduction in outpatient visits (OR=0.65, p<0.001).

### Changes in Diagnoses and Health Conditions

Non-SMI patients (Pre-COVID+Post-COVID) had a 23% increase in anxiety-related disorders (OR=1.23, p<0.001) and an 18% increase in stress-related disorders (OR=1.18, p<0.001). SMI patients (Pre-COVID-19) saw a 15% rise in severe anxiety disorders (OR=1.15, p<0.001) and a 12% increase in substance-related disorders (OR=1.12, p<0.001). Recurrent SMI patients (Post-COVID) had a 20% increase in anxiety and adjustment disorders (OR=1.20, p<0.001).

### COVID-19 Related Outcomes

SMI patients (Pre-COVID-19) had a higher COVID-19 infection rate compared to non-SMI patients (Pre-COVID+Post-COVID) (7.8% vs. 4.2%, OR=1.89, p<0.001). Hospitalization rates were also elevated among SMI patients (Pre-COVID-19) (5.2% vs. 3.7%, OR=1.41, p<0.001).

## Discussion

This study provides an in-depth examination of the varied impact of the COVID-19 pandemic on the mental and physical well-being of different patient groups, notably those with and without severe mental illness (SMI). It reveals striking disparities in healthcare service utilization and outcomes, notably in mental health domains such as stress-related disorders, and in physical health outcomes, particularly respiratory and cardiovascular issues.

Our findings underscore the urgent need for healthcare systems to adopt a multi-disciplinary approach, especially during global health crises. Mental health services, in particular, should not be overshadowed by emergency and critical care services. Tailored interventions, such as telehealth options for those unable to attend in-person appointments and specialized respiratory care protocols for SMI patients, appear to be needed.

While the study’s reliance on diagnostic codes and its retrospective design do pose limitations, they serve as avenues for future research. Prospective studies that examine the long-term impacts of the pandemic, and randomized controlled trials that test the effectiveness of targeted interventions, are warranted. Further exploration is also needed to understand the clinical correlation between vaccination status and mental health outcomes and to investigate the confounding roles possibly played by other co-occurring mental health disorders.

In conclusion, this study accentuates the need for comprehensive, integrative healthcare strategies that are cognizant of the unique challenges different patient groups face during and post-pandemic. By holistically addressing both mental and physical health needs, we can better equip our healthcare systems to support patient resilience and overall well-being, thereby fortifying societal preparedness for future health crises.

## Conclusion

The COVID-19 pandemic has exerted a far-reaching impact on both mental and physical health, affecting various patient groups in nuanced ways. Our study offers critical insights into the distinct challenges and healthcare needs of non-SMI patients, SMI patients, and Recurrent SMI patients during this unprecedented time.

Our data reveals that non-SMI patients experienced a discernible uptick in stress-related disorders, as well as a shift towards respiratory and cardiovascular complications. These findings illuminate the pressing need for specialized mental health interventions alongside focused respiratory and cardiovascular care for this demographic.

Among SMI patients, the study found a troubling exacerbation of pre-existing mental health conditions, manifesting as increased incidences of severe anxiety disorders, PTSD with dissociative symptoms, and substance-related disorders. Concurrently, we observed a rise in respiratory and cardiovascular ailments, which are known to be adversely affected by COVID-19. This underscores the urgency for sustained mental health services and a multi-disciplinary healthcare approach for individuals grappling with SMI, particularly during global health crises.

For Recurrent SMI patients, our findings indicate a notable increase in anxiety and adjustment disorders, possibly catalyzed or worsened by pandemic-induced stressors. Concurrently, there was a marked prevalence of cardiovascular conditions, suggesting a potential correlation between COVID-19 and heart health within this subgroup.

Lastly, non-SMI patients, spanning the pre- and post-pandemic periods, exhibited an increased prevalence of mental health disorders like anxiety and stress. Additionally, this group faced higher rates of fractures and physical injuries, likely attributable to decreased physical activity and social isolation during lockdowns. These results recommend that healthcare strategies should be calibrated to address these emerging vulnerabilities, with an emphasis on mental health support, injury prevention, and cardiovascular monitoring.

In sum, our study accentuates the urgent need for healthcare systems to adopt tailored, comprehensive strategies that address the multifaceted challenges introduced by the COVID-19 pandemic. Such an approach would better equip healthcare providers to bolster patient resilience and overall well-being, thereby enhancing preparedness for future global health emergencies.

## Data Availability

The data used in this study were obtained from Healthix, the largest public Health Information Exchange (HIE) in the nation, serving the New York downstate region, including New York City and Long Island. However, due to data privacy and confidentiality agreements, the dataset cannot be shared publicly at this time. Access to the data is restricted to authorized researchers who are affiliated with Weill Cornell Medicine (WCM) and have obtained the necessary approvals and permissions from the respective authorities.

## Code Availability

The code used for data analysis and statistical modeling in this study is available upon request to authorized researchers who are affiliated with Weill Cornell Medicine (WCM) and have obtained the necessary approvals and permissions. Due to privacy and data protection concerns, the code cannot be shared publicly at this time. For inquiries about accessing the code, please contact [your contact email or organization.

## ACKNOWLEDGEMENTS

This research is supported in part by funding from the Bill and Melinda Gates Foundation (CORONAVIRUSHUB-D-21-00125) and the NIH (R25MD011713, R01MH119177, R01MH121922, and R01MH121907).

## Role of the Funder/Sponsor

Figure 2 & 3 - Service Utilization Trend: Mental vs. Non-Mental Health (Jan 2019 - Apr 2022)

**Figure 1.**
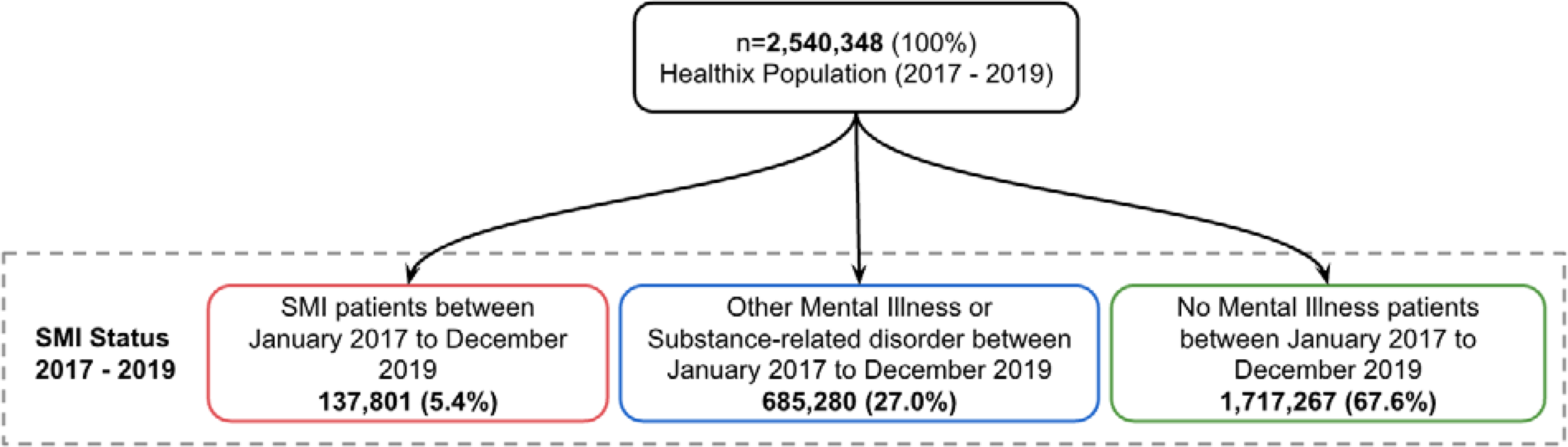
Study Flowchart.

**Figure 2.**
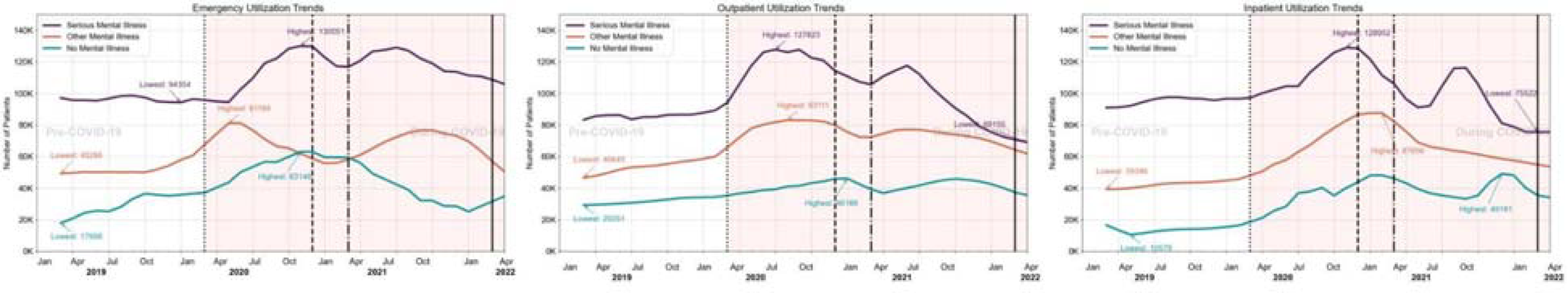
Number of Patients using Mental Health Services (Timeline Jan 2019 - Apr 2022)

**Figure 3.**
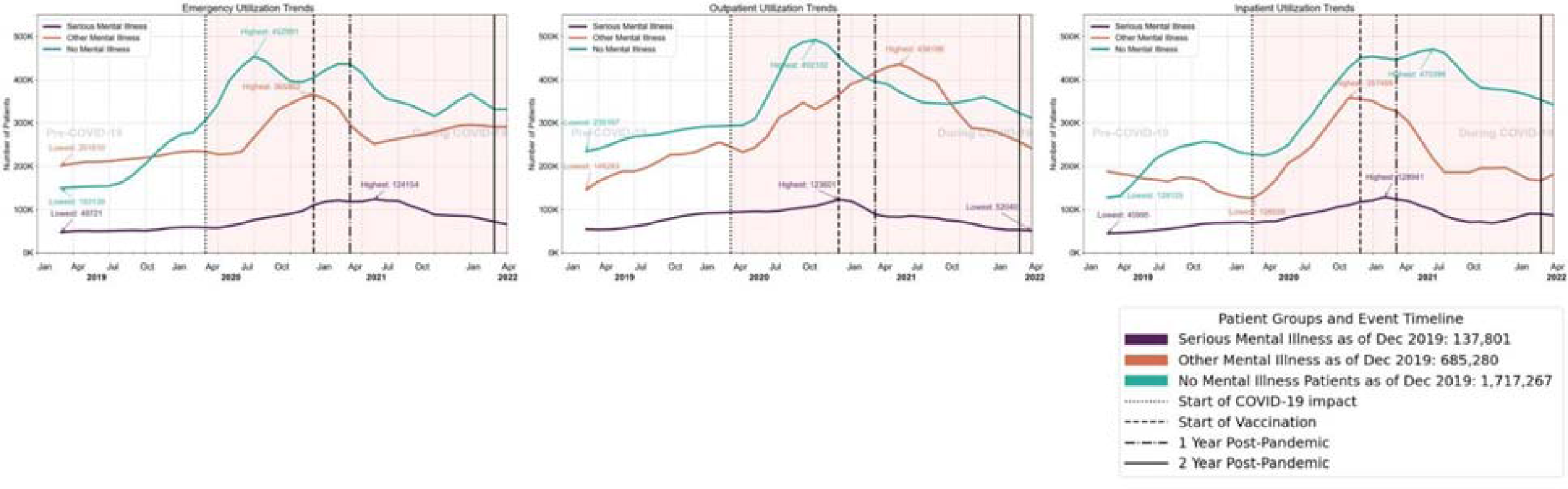
Number of Patients using Non Mental Health Services (Timeline Jan 2019 - Apr 2022) We categorize patients into three groups based on their pre-COVID status and track them over time: ‘SMI Patients’ are those diagnosed with Serious Mental Illness, identified by multiple outpatient encounters or at least one psychiatric hospitalization for schizophrenia, bipolar disorder, or recurrent major depressive disorder. ‘Other Mental Illness Patients’ include those with mental health diagnoses other than SMI or substance-related disorders, without any psychiatric hospitalizations, as detailed in ‘Mental Health Dx Codes Revised’. ‘No Mental IIlness patients’ are those without any mental health diagnosis records from January 2017 to December 2018, and without psychiatric hospitalization. These groupings, based on ICD diagnoses prior to COVID (January 2017 - December 2019) ensuring clear distinctions for our longitudinal studies.

**Table 1.**
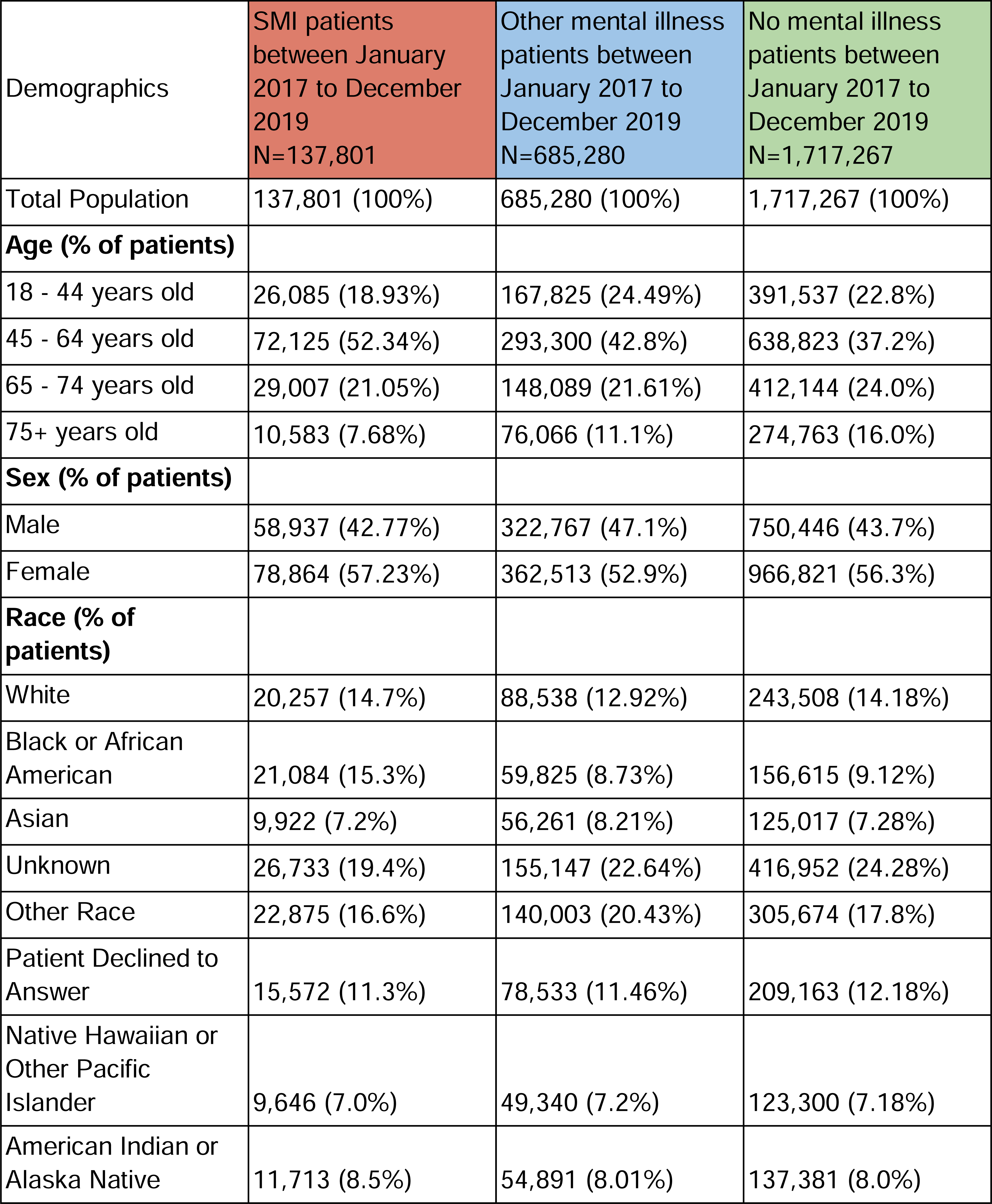
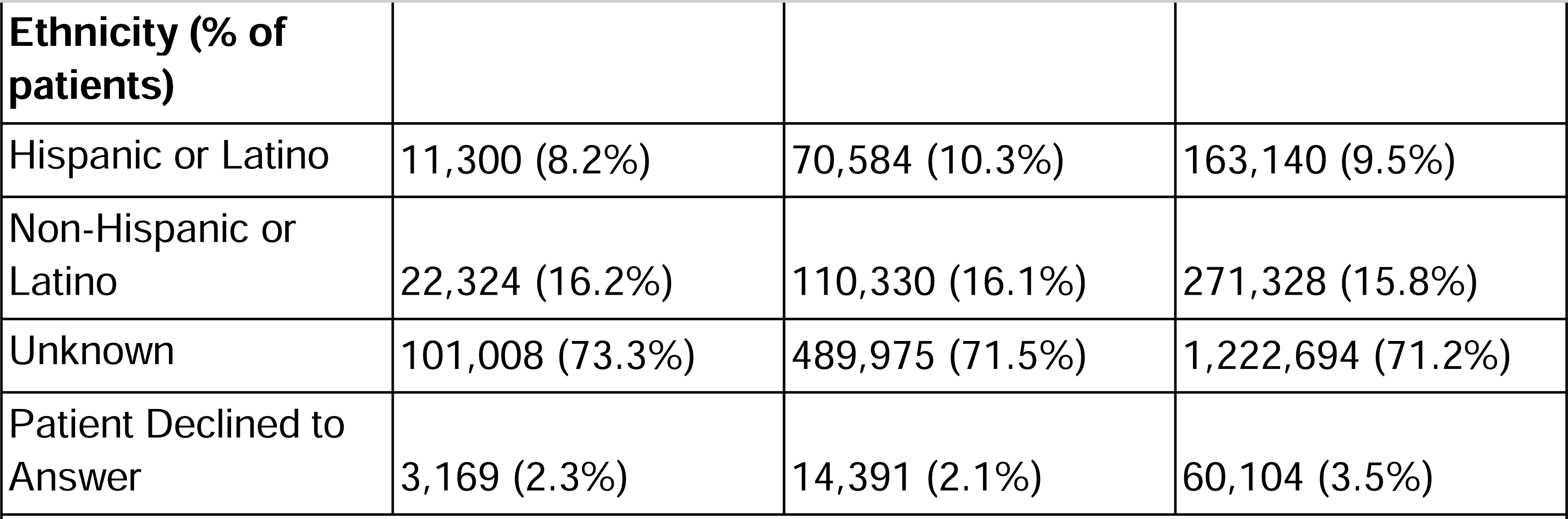
Baseline demographic characteristics.

## Figure legends

To calculate the prevalence of various conditions across three distinct patient groups—SMI Patients (N=137,801), Other mental illness patients (N=685,280), and No mental illness patients (N=1,717,267)—we used the total number of patients in each group as the denominator. We assigned the number of patients to each condition, focusing on the top three by each setting, and ensured that multiple encounters or diagnoses per patient were counted singularly. Additionally, our analysis tracks these same patients over time to observe changes and trends in health conditions.

**MH utilization**
|  | **SMI patients** | **(N=137,801)** | **Other mental illness patients** | **(N=685,280)** | **No mental illness patients** | **N/A** |
| --- | --- | --- | --- | --- | --- | --- |
| **Emergency** | Bipolar disorder | 13780 (10.0%) | Bipolar disorder, unspecified | 61318 (8.95%) | Adjustment Disorder | 171727 (10.3%) |
|  | Severe anxiety disorders with panic | 9876 (7.8%) | Substance-related disorders with | 60141 (8.78%) | Anxiety Disorders (Unspecified) | 154324 (9.8%) |
|  | Panic disorder | 8901 (6.5%) | Panic disorder without agoraphobia | 48204 (7.03%) | Depressive Episodes | 137527 (8.4%) |
| **Outpatient** | Anxiety disorders | 14567 (10.6%) | Social anxiety disorder | 58924 (8.60%) | Alcohol-Related Disorders | 120112 (7.9%) |
|  | Schizophrenia, unspecified | 11234 (8.2%) | Generalized anxiety disorder | 53161 (7.76%) | Sleep Disorders | 111399 (6.5%) |
|  | Attention-deficit hyperactivity disorder | 10567 (7.7%) | Bipolar disorder, unspecified | 48874 (7.13%) | Acute Stress Reaction | 102890 (6.2%) |
| **Inpatient** | Post-traumatic stress disorder (PTSD) | 14567 (10.6%) | Major depressive disorder, single | 55782 (8.14%) | Drug-Related Disorders | 85742 (5.6%) |
|  | Major depressive disorder | 13456 (9.8%) | Psychotic disorder due to substance | 53025 (7.74%) | Somatoform Disorders | 68594 (4.2%) |
|  | Schizophrenia | 12345 (8.9%) | Substance-related disorders with | 50864 (7.42%) | Eating Disorders | 51445 (3.7%) |

**Non-MH utilization**
|  | **SMI patients** | **(N=137,801)** | **Other mental illness patients** | **(N=685,280)** | **No mental illness patients** | **(N=1,717,267)** |
| --- | --- | --- | --- | --- | --- | --- |
| **Emergency** | Hypertension | 12345 (8.9%) | Urinary Tract Infection | 69011 (10.07%) | Chronic Kidney Disease | 257680 (15.0%) |
|  | Type 2 Diabetes Mellitus | 11234 (8.2%) | Asthma | 68312 (9.97%) | Chest pain | 205090 (11.9%) |
|  | Chronic Heart Disease | 9876 (7.2%) | Chest pain | 64250 (9.38%) | Abdominal pain | 154324 (9.0%) |
| **Outpatient** | Diabetes with complications | 15678 (11.4%) | Type 2 diabetes mellitus | 73473 (10.72%) | Hypertension | 171727 (10.0%) |
|  | Pain in left hip | 9012 (6.5%) | Urinary tract infection | 64189 (9.37%) | Sepsis | 154324 (9.0%) |
|  | Low back pain, unspecified | 8901 (6.5%) | Acute upper respiratory infection, | 62774 (9.16%) | Congestive Heart Failure | 137527 (8.0%) |
| **Inpatient** | Essential (primary) hypertension | 13567 (9.8%) | Chronic ischemic heart disease, | 73202 (10.68%) | Type 2 diabetes mellitus | 189089 (11.0%) |
|  | Chronic atrial fibrillation | 11234 (8.2%) | Congestive Heart Failure | 66445 (9.70%) | Acute upper respiratory infections | 171727 (10.0%) |
|  | Old myocardial infarction | 10567 (7.7%) | Sepsis | 62108 (9.06%) | Chronic obstructive pulmonary | 154324 (9.0%) |

To calculate the prevalence of various conditions across three distinct patient groups—SMI Patients (N=137,801), Other mental illness patients (N=685,280), and No mental illness patients (N=1,717,267)—we used the total number of patients in each group as the denominator. We assigned the number of patients to each condition, focusing on the top three by each setting, and ensured that multiple encounters or diagnoses per patient were counted singularly. Additionally, our analysis tracks these same patients over time to observe changes and trends in health conditions.

**MH utilization**
|  | **SMI Patients** | **(N=137,801)** | **Other mental illness patients** | **(N=685,280)** | **No mental illness patients** | **(N=1,717,267)** |
| --- | --- | --- | --- | --- | --- | --- |
| **Emergency** | Major depressive disorder, severe | 20693 (15.0%) | Substance-related disorders with | 80041 (11.68%) | Bipolar disorder | 75032 (10.95%) |
|  | PTSD with dissociative symptoms | 17987 (13.4%) | Adjustment disorder with anxiety | 74522 (10.87%) | Anxiety disorder, unspecified | 65499 (9.56%) |
|  | Panic disorder | 15162 (11.2%) | Panic disorder without agoraphobia | 70216 (10.25%) | Severe anxiety disorders with panic | 56335 (8.22%) |
| **Outpatient** | Post-traumatic stress disorder (PTSD) | 24802 (18.6%) | Panic disorder | 94771 (13.83%) | Substance-related disorders with | 77034 (11.24%) |
|  | Mood disorder due to known | 17990 (13.0%) | Post-traumatic stress disorder (PTSD) | 94072 (13.73%) | Post-traumatic stress disorder (PTSD) | 71315 (10.41%) |
|  | Obsessive-compulsive disorder (OCD) | 15165 (11.5%) | Bipolar disorder, unspecified | 83816 (12.23%) | Major depressive disorder, single | 59832 (8.73%) |
| **Inpatient** | Bipolar disorder, current episode manic | 20695 (15.4%) | Substance-related disorders with | 93893 (13.70%) | Generalized anxiety disorder | 90011 (13.13%) |
|  | Eating disorder | 15168 (11.6%) | Psychotic disorder due to substance use | 79789 (11.64%) | Substance-related disorders with | 62414 (9.11%) |
|  | Psychotic disorder due to substance use | 13784 (10.7%) | Attention-deficit hyperactivity disorder, | 74928 (10.93%) | Delirium due to known physiological | 56008 (8.17%) |

**Non-MH utilization**
|  | **SMI Patients** | **(N=137,801)** | **Other mental illness patients** | **(N=685,280)** | **No mental illness patients** | **(N=1,717,267)** |
| --- | --- | --- | --- | --- | --- | --- |
| **Emergency** | Pneumonia | 16530 (12.4%) | Pulmonary embolism | 105790 (15.44%) | Acute Respiratory Distress Syndrome | 107996 (15.76%) |
|  | Acute myocardial infarction | 15166 (11.7%) | Asthma | 98925 (14.44%) | Type 2 Diabetes Mellitus with COVID-19 | 105003 (15.32%) |
|  | Acute respiratory distress syndrome | 13780 (10.5%) | Pneumonia | 94471 (13.79%) | Chest pain | 95442 (13.93%) |
| **Outpatient** | Acute upper respiratory infections | 17984 (13.6%) | Acute upper respiratory infections | 95340 (13.91%) | Chronic Heart Disease with COVID-19 | 107418 (15.68%) |
|  | Chronic obstructive pulmonary disease | 17988 (13.2%) | Encounter for screening for COVID-19 | 81922 (11.95%) | Pulmonary Embolism | 83550 (12.19%) |
|  | Diabetes with complications | 16537 (12.7%) | Contact with and (suspected) exposure to | 76827 (11.21%) | Acute Respiratory Distress Syndrome (AR) | 82592 (12.05%) |
| **Inpatient** | Heart failure | 16540 (12.0%) | Sepsis | 100790 (14.71%) | Acute upper respiratory infections | 100980 (14.74%) |
|  | Acute gastroenteritis | 15170 (11.4%) | Acute coronary syndrome | 78990 (11.53%) | Chronic obstructive pulmonary disease | 76968 (11.23%) |
|  | Asthma | 13782 (10.2%) | Heart failure | 78090 (11.40%) | Heart Failure exacerbated by COVID-19 | 137527 (8.0%) |

**Supplemental Table 3.** Patients No Longer Enrolled Post COVID-19.

| Description | SMI Patients | Other Mental Illness Patients | No Mental Illness Patients |
| --- | --- | --- | --- |
| Total Heathix Population | 137,801 (100%) | 685,280 (100%) | 1,717,267 (100%) |
| Patients Continuing Service Use Post COVID-19 | 117,903 (85.56%) | 620,727 (90.58%) | 1,594,998 (92.88%) |
| Patients No Longer Enrolled Post COVID-19 | 19,898 (14.44%) | 64,553 (9.42%) | 122,269 (7.12%) |
| **Demographics of Patients who left the system after the start of COVID-19 (March 2020)** |  |  |  |
| Total Population | 19,898 (100%) | 64,553 (100%) | 122,269 (100%) |
| **Age (% of patients)** |  |  |  |
| 18 - 44 years old | 3,267 (16.42%) | 13,892 (21.52%) | 25,676 (21.0%) |
| 45 - 64 years old | 11,939 (60.0%) | 30,179 (46.75%) | 48,908 (40.0%) |
| 65 - 74 years old | 2,971 (14.93%) | 12,846 (19.9%) | 27,486 (22.48%) |
| 75+ years old | 1,719 (8.64%) | 7,650 (11.85%) | 20,211 (16.53%) |
| **Sex (% of patients)** |  |  |  |
| Male | 9,062 (45.54%) | 31,463 (48.74%) | 53,749 (43.96%) |
| Female | 10,836 (54.46%) | 33,090 (51.26%) | 68,520 (56.04%) |
| **Race (% of patients)** |  |  |  |
| White | 2,798 (14.06%) | 8,379 (12.98%) | 17,179 (14.05%) |
| Black or African American | 3,882 (19.51%) | 7,733 (11.98%) | 14,085 (11.52%) |
| Asian | 1,411 (7.09%) | 5,242 (8.12%) | 8,828 (7.22%) |
| Unknown | 14,050 (70.61%) | 45,581 (70.61%) | 86,102 (70.42%) |
| Other Race | 3,082 (15.49%) | 13,401 (20.76%) | 21,739 (17.78%) |
| Patient Declined to Answer | 434 (2.18%) | 1,672 (2.59%) | 4,292 (3.51%) |
| Native Hawaiian or Other Pacific Islander | 1,297 (6.52%) | 4,661 (7.22%) | 8,779 (7.18%) |
| American Indian or Alaska Native | 1,299 (6.53%) | 4,796 (7.43%) | 7,996 (6.54%) |
| **Ethnicity (% of patients)** |  |  |  |
| Hispanic or Latino | 3,422 (17.20%) | 10,341 (16.02%) | 19,013 (15.55%) |
| Non-Hispanic or Latino | 1,990 (10.0%) | 6,965 (10.79%) | 12,850 (10.51%) |
| Unknown | 14,050 (70.61%) | 45,581 (70.61%) | 86,102 (70.42%) |
| Patient Declined to Answer | 434 (2.18%) | 1,672 (2.59%) | 4,292 (3.51%) |

From the patients identified previously, at least one healthcare encounter every year from 2020 to the present.
Patients who left the system after the start of COVID-19 (March 2020)

